# ‘How can I explain it to my daughter if I don’t know?’: Understanding health communication and routine and HPV vaccine hesitancy in rural Indigenous communities of Guatemala

**DOI:** 10.1101/2025.08.01.25332635

**Authors:** Nadine Ann Skinner, Lucia Abascal-Miguel, Emily Lopez, Magda Sotz, Julia Coxaj, Victoria Ward, Jamie Sewan Johnston, Nadia Diamond-Smith, Anne Kraemer Diaz

**Author notes:** Corresponding author Nadine Ann Skinner.

## Abstract

Guatemala saw a decline in childhood immunisation and HPV vaccine rates even before the COVID-19 pandemic. Indigenous communities, which make up over 40% of the population, face especially low vaccination rates. We sought to understand how to address childhood and HPV vaccine hesitancy to increase uptake among Indigenous communities in rural Guatemala. Using a community-engaged design process, we held (1) 9 Focus Group Discussions (FGDs) with 41 Indigenous community members in 11 communities in 3 departments and (2) 16 In-Depth Interviews (IDIs) with health workers. Based on participants’ preferences, FGDs/IDIs were conducted in Spanish, K’iche, or Kaqchikel. Community members and health workers indicated individual, interpersonal, and systemic barriers to vaccination within rural Indigenous communities in Guatemala, including limited knowledge, rumours, and a need for improved health worker knowledge and skills. Despite the identified barriers to vaccine uptake, community members and health workers believed that attitude shifts and enhanced access to health information and technology would improve health and lead to improved vaccination rates. Future vaccine campaigns must focus on targeted communication strategies that address the specific beliefs, rumours, lack of community information and health worker knowledge, and reliance on community networks for health and vaccine information.

## Introduction

Since 2017, childhood immunisation rates in Guatemala have declined, with coverage for the third dose of the diphtheria, pertussis, and tetanus (DPT) vaccine dropping from 91% (2017) to 79% (2022) and the second dose of the measles vaccine falling from 89% to 69% by 2022 [1]. Human papillomavirus (HPV) vaccine coverage in Guatemala has also declined from 32% of girls completing vaccination in 2018 to just 18% in 2022, which is well below the 90% coverage goal set by the Pan American Health Organization (PAHO) and the World Health Organization (WHO) [1,2].

Almost all cervical cancer is linked to infection with HPV [2–5]. In Guatemala, cervical cancer is the second most prevalent and has the second highest mortality risk among cancers affecting women [3]. HPV vaccines are highly effective preventative strategies against HPV infection and cervical cancer, have excellent safety profiles, and are the WHO’s primary strategy to eliminate cervical cancer [2,6]. Despite the proven efficacy of vaccines, disparities in vaccine coverage persist, affecting both childhood immunisations and HPV vaccinations.

Over 40% of Guatemala’s population identifies as Indigenous, and Indigenous children have persistent and pronounced health disparities and higher under-five mortality in low-income and rural areas [7–9]. Outside of Guatemala City, areas with the lowest vaccination rates often have the highest Indigenous populations [8,10]. Indigenous rural women in Guatemala have lower HPV awareness, lower cervical cancer screening rates, and receive less follow-up care [11–15]. This experience reflects high global rates of infectious diseases and poor access to health services for Indigenous populations[16]. HPV infection rates are notably higher among Indigenous populations compared to non-Indigenous groups [13,17–18], which leads to disproportionate cervical cancer incidence and mortality rates for Indigenous women [13,17].

Barriers to vaccine uptake are multifaceted. Practical issues, such as affordability, ease of access, service quality, geography, needing permission to visit a doctor, or reluctance to visit clinics alone [19] may create barriers. A study at four Guatemalan public clinics indicated that vaccine shortages due to political instability, distance, travel costs, and wait times may have contributed to delayed vaccine timing and completion rates [20]. However, understanding vaccine barriers goes beyond practical issues and requires addressing knowledge and beliefs about vaccine-preventable diseases, perceived risks, and confidence in vaccines. Confidence, in this context, refers to trust in the efficacy and safety of vaccines, health services, and the motivations of health policymakers [17]. In Latin America and the Caribbean (LAC), attitudes toward vaccination impede uptake regardless of vaccine type [22–23].

Limited awareness about vaccine availability and effectiveness and insufficient information from health authorities and healthcare personnel can fuel misconceptions about vaccines, lead individuals to view vaccination as unnecessary [21], and reduce parental intent to vaccinate [22]. In previous research in Guatemala, parents expressed concerns over the perceived excessive number of vaccines, and for the HPV vaccine, they shared misinformation, perceptions of the vaccine as ‘experimental,’ incorrect or insufficient information regarding adverse effects and efficacy, and fears it may encourage early sexual activity in daughters [22,24].

Throughout the LAC region mistrust in health systems, lack of physician recommendations, and social pressures contribute to vaccine hesitancy [22]. Vaccine hesitancy refers to the delay or refusal of vaccination despite its availability [21]. Additionally, Indigenous women face barriers, such as discrimination, language differences, and distrust of healthcare providers, further limiting access to vaccination services [11,17]. With vaccine coverage below global targets, especially in Indigenous communities, this study aimed to identify barriers to vaccination and health information needs in Indigenous communities in rural Guatemala.

## Methods

### Study design

Using a community-engaged design process, we conducted individual in-depth interviews (IDI) and focus group discussions (FGD) about immunisation barriers, misinformation or lack of vaccine information, individual and group beliefs about vaccines, and reasons for vaccine hesitancy in eleven Indigenous Maya communities in three Guatemalan departments.

The research team developed structured guides for the IDIs and FGDs in English, which were then reviewed and revised. The guides were translated into Spanish and piloted with four community members and two health workers. After the pilot, guides were further revised and translated into K’iche or Kaqchikel, the two most common Indigenous Mayan languages in Guatemala, including in the areas where the study took place [8]. Full English copies of the guides are included in the appendix.

The IDI guide has five sections of questions about health workers’ experiences administering childhood and HPV vaccines, responding to community vaccine questions, receiving information and training about vaccines, and their responses to potential educational materials being developed. Using a snowball technique, Wuqu’ Kawoq’s staff members and interviewers recruited health workers into the study in July 2023. Four interviewers first obtained verbal consent from 16 health workers, who then completed the interview followed by a demographic survey. The IDIs took place over Zoom, and health workers were encouraged to find a quiet, private place to participate. The one-hour long IDIs conducted in Spanish were automatically transcribed via Zoom’s transcription software, checked against the audio recording, and revised for accuracy. The transcripts were translated into English with the support of automated translation tools and reviewed for accuracy by bilingual team members.

The FGD guide has six sections about community members’ beliefs, attitudes, knowledge, and behaviours with childhood immunisations and the HPV vaccine. The guide also explored community members’ trusted sources for health information and probed responses to potential materials being developed. The 41 community members were recruited in August 2023 to participate in the FGDs by health workers and through a snowball technique to engage participation. After obtaining verbal consent with each participant, one interviewer led the nine one-hour long FGDs, which took place in quiet, private rooms. After each FGD, the interviewer transcribed the FGDs and translated them from K’iche or Kaqchikel into Spanish. Bilingual researchers translated the transcripts into English.

### Study population

The research took place in 11 Indigenous Maya communities (82.1%-98.8% Maya) [8] in three priority departments identified by the Ministry of Public Health and Welfare. These include three communities in Sololá: 1) San Andres Semetabaj, 2) Sololá, 3) San Lucas Tolimán; six in Chimaltenango: 4) Patzún, 5) Patzicía, 6) Tecpán, 7) San Juan Comalapa, 8) Santa Apolonia, 9) San Martín Jilotepeque; and two in Sacatepéquez: 10) Santiago and 11) Santa María de Jesús. Characteristics of the focus group and interview participants can be found in the following tables.

**Table 1.** Focus group discussion population characteristics (N=40)

|  |  |  |
| --- | --- | --- |
| Department | Chimaltenango<br>Sacatepéquez<br>Sololá<br>Other | 60%<br>28%<br>13%<br>0% |
| Rural/Urban | Urban<br>Semi-urban<br>Rural | 78%<br>0%<br>23% |
| Gender | Female<br>Male | 98%<br>3% |
| Age | 18-24<br>25-34<br>35-44<br>45-60<br>Older than 60 years | 8%<br>30%<br>38%<br>23%<br>3% |
| Primary Language | Spanish<br>K'iche<br>Kaqchikel | 48%<br>13%<br>40% |
| Caregiver Role* | Mother<br>Father<br>Grandparent | 95%<br>5%<br>3% |
| Number of Children | 1 child<br>2-3 children<br>4-5 children<br>6-7 children | 33%<br>35%<br>28%<br>5% |

\*Note: Respondents could list more than one type of caregiver role, therefore responses add up to more than 100%. One community member did not complete a demographic survey after the focus group. Rounding may result in values greater than 100%.

**Table 2.**
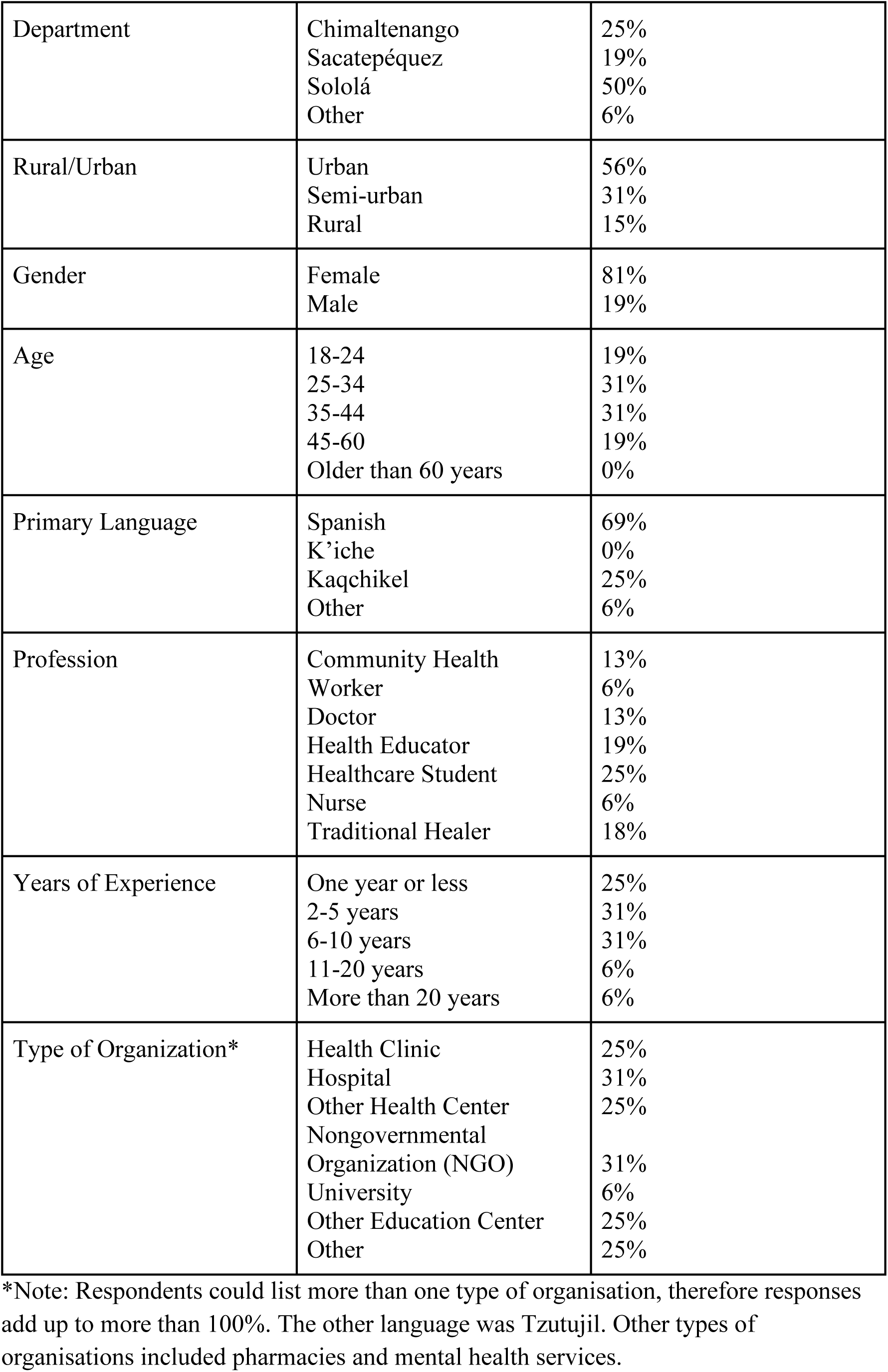
In depth interview population characteristics (N=16)

### Community-engaged approach

This study applied a community-engaged education design, grounded in human-centred design principles [25–26], to explore vaccine perceptions in Indigenous Maya communities. Indigenous health workers and community members were engaged from the start in two feedback sessions. Their input led to expanding the research questions to include a broader range of vaccines, including HPV. Community input also shaped the research instruments, with adjustments made to reflect local terminology and cultural nuances, particularly concerning HPV vaccine discussions. Study findings were shared with community health workers for further feedback and to create education videos and infographics in Spanish, K’iche, and Kaqchikel, which are being disseminated in Guatemala.

### Analysis

Based on a preliminary literature review and community input, two team members conducted a rapid analysis, generating initial themes that were reviewed by the full research team. We then revised the codebook and comparative analysis plan to evaluate ideas about access, attitudes, knowledge, and behaviours. Deductive codes derived from our research questions focused on access to healthcare, attitudes about vaccination, and sources of information. Inductive codes from the rapid analysis included shifting vaccine attitudes, availability of vaccine information, and the impact of the COVID-19 vaccine on overall hesitancy.

Two researchers coded each transcript in Dedoose, with a third researcher conducting a final reliability check. We used automatic upcoding, starting at the most basic level of the code applicable, before applying higher-level coding. The entire sentence or paragraph was coded to capture the full context and meaning. Code definitions allowed for the application of synonyms or implicit codes. The exception is two ‘behaviours’ codes, ‘community behaviours with childhood vaccines’ and ‘community behaviours with HPV’, which were only applied when it was explicitly stated whether community members vaccinated or did not vaccinate their children. Any questions, uncertainties, or discrepancies that arose during the coding process were documented, discussed, and resolved in review meetings between each round. Code applications were then analysed for primary themes. Once consensus was reached on the primary themes they were presented to local interviewers for further validation.

### Author reflexivity statement

The all female research team includes four researchers in Guatemala and five affiliated with U.S. institutions. These researchers bring diverse backgrounds in qualitative methods, health education, clinical practice, and health research. We actively practised intrapersonal reflexivity to ensure relevance, equitable treatment, and ethical engagement with Indigenous communities in weekly discussions and reflections on the research process, focusing on refining questions, analyses, and findings. We are dedicated to Wuqu’ Kawoq’s commitment to healthcare research led by Indigenous health workers. FGDs and IDIs were conducted in Spanish, K’iche, or Kaqchikel by researchers trained in qualitative interview methods. FGDs were facilitated by an Indigenous Maya interviewer, who met with community members in person to ensure trust, safety, and inclusion were built into the research process. Multilingual staff members focused on contextual reflexivity by reviewing and revising transcripts and instruments to ensure that translations were accurate and captured cultural nuance.

## Results

### Immunisations and HPV vaccination concerns and fears

Across the focus groups and interviews, community members and health workers shared that some had fears of vaccination and refused vaccines. One community member shared ‘there are families that do not agree with vaccinations… They used to knock on doors to offer vaccinations… they always told them, I don’t have children’ FGD 001. Another community member stated ‘We have a neighbour who is older than us, she hides her children from the vaccine. The nurses there are reaching out and the lady never comes out… The girls are not vaccinated’ FGD 002. Health workers shared:

> ‘Some parents remain extremely reluctant. We have to visit their homes to look for them. Some hide, they stay indoors because they disagree with vaccinating their children. It’s challenging to engage with them, you don’t have a space to address their doubts’ IDI 009.

Families primarily expressed fears of side effects, particularly fever, and some believed these risks outweighed the benefits. This belief was often combined with a limited understanding of vaccine purpose. A community member shared ‘fevers make [children] very upset. It’s worrying because, in their little head, they feel very sick, very bad. You don’t know what is going to happen to them, because the fever is not a game’ FGD 009. Health workers shared this concern was common:

> ‘There are parents who are reluctant and don’t want to vaccinate their children… They don’t want to accept the vaccine mostly because of the side effects. They know that they’re going to get a fever… and they think that the vaccine is not necessary’ IDI 007.

Community members also expressed concern about injection site swelling, particularly after the second DPT dose. Several community members feared swelling could lead to paralysis if not properly managed. One community member shared:

> “We went to visit a sick child… we asked the mother what had happened, why the girl couldn’t walk. The mother told us, “… I would tell you not to give [children] the four-year vaccine, because my little girl was left like this after she was vaccinated.” She can’t walk… I imagine the fault here was with the person who administered it, meaning it was not done in the place where it was supposed to be…’ FGD 001.

In another focus group, two community members discussed their concerns:

> Community member 1: ‘The one that has me concerned is the vaccine for four-year-olds… some people say that the children have not been able to move their feet and have been paralyzed’.

> Community member 2: ‘I have concerns about the vaccine because my baby will be four-years-old next year. Both my husband and I are unsure about it. We don’t know if this vaccine will affect him’ FGD 009.

Awareness of the HPV vaccine was limited, but across interviews and focus groups there were fears that it causes sterility in girls. One community member said, ‘some girls and some mothers do not accept this vaccine either because someone has told them that this vaccine is to sterilise them’ FGD 001 and another said, ‘there are those who fear that the vaccine might be aimed at sterilising them so that they can have unrestricted sexual activity’ FGD 006. Some were confused, ‘I really don’t know how true it is… they say they leave you sterile… that’s my doubt’ FGD 004 and others were convinced ‘this vaccine prevents girls from having more children’ FGD 008. Health workers also heard this belief ‘I have heard people say I am not going to vaccinate my daughter because the vaccine can sterilise’ IDI 007. Another shared:

> ‘… we had challenges with the promotion and coverage of the HPV vaccine… When we offered the vaccine in the community, they refused it, they didn’t want it because they said that the vaccine would make them sterile. Many mothers were against it and did not want to give it to their children’ IDI 012.

### Absence of information

Health workers and community members cited the lack of accessible vaccine information as a primary main challenge. Community members knew various vaccines were available but often lacked an understanding of the specific purposes and benefits of each vaccine, which related to misinformation and families to not fully grasping the associated benefits or risks of vaccination. A community member shared:

> ‘One of my relatives did not want to give a vaccine to her son because she did not know what the vaccine was for… She even had a disease, hepatitis, mainly because she did not give him that vaccine. So, it was not until later that one realises what the benefits of a vaccine are’ FGD 005.

Another community member expressed their frustration stating ‘More than everything, more information is needed. More information about vaccines, what each vaccine is for’ FGD 006. Health workers shared the concern, stating:

> ‘We have to vaccinate children against measles… What are the measles? Or what are the consequences to the child if they don’t get vaccinated against measles? A lot of times they don’t understand why each vaccine… we do know that vaccines are important, but we are not very clear about what diseases each vaccine is preventing’ IDI 007.

There was a lack of information about the HPV and its vaccine. One community member stated:

> ‘In my understanding, it’s [HPV is] related to sexual activity… If I had classes or more information, I would learn there… How can I explain it to my daughter if I don’t know?’ FGD 006.

Health workers also expressed frustration with the limited information available about HPV and the vaccine, and some were even unaware of the vaccine’s existence. One health worker shared:

> ‘the community often fails to connect the vaccine to its primary goal of preventing uterine cancer… This lack of understanding contributes to hesitancy and reluctance among community members’ IDI 009.

Another stated:

> ‘In the community in general they are totally unaware of this vaccine… The leaders ask what it means, HPV, because one mentions it, but they really don’t know the meaning of each word, why they have to get vaccinated, or what is the benefit…. to have clear information about the function of this vaccine… the community does not really know that it is a risk’ IDI 015.

### Health worker training, communication, and community trust

Some health workers noted insufficient education and training as contributing to the lack of vaccine information being shared and poor healthcare practices. One health worker shared her experience ‘When I was in health in the municipality, I didn’t hear about [the HPV vaccine]’ IDI 003 and another said ‘we don’t have a space to be able to train… we really lack enough information to be able to respond when we are asked [about the HPV vaccine]’ IDI 015. Several health workers also indicated they did not know how to speak to community members about the HPV vaccine given the sexual transmission of HPV and cultural taboos about discussing sex. One health worker shared ‘when we mention it’s sexually transmitted, it becomes a taboo topic for some communities to discuss’ IDI 009.

Community members also shared negative experiences with health workers and times when they not providing vaccine information. One mother shared:

> ‘I have my son, the oldest, and the nurse scolded him. She said that he is a man, he has to be brave. So, I corrected her. I told her, he is brave but the vaccination hurts… I will say that many people out of respect, out of pity, out of fear, for whatever reason, do not correct the nurses, or the nurses are very aggressive with the children, and that generates trauma for them. My son, the older one, is terrified of injections… I stopped taking him to get the vaccines’ FGD 001.

Another community member shared ‘Sometimes the mothers don’t want to go because they were given bad attention. [The nurses] even scold them’ FGD 008.

Parents were upset when vaccines were given without an explanation, and these experiences would increase their distrust of vaccinations and the health system. One health worker shared:

> ‘I have seen nurses taking the baby, giving them all the vaccines, telling them that they are going to react and that’s it, time for the next child… they don’t take the time to explain, look this is for your measles, this is for rotavirus… this is going to help your child not to get a lot of diarrhoea’ IDI 009.

Another health worker shared ‘sometimes they [families] have had bad experiences with vaccines. There’s misapplication, misinformation… in some communities that I have visited, they tell me that they force them to vaccinate’ IDI 011.

However, community members and health workers also noted that health centres were the primary source of vaccine information, and community health workers play a crucial role in disseminating vaccine information through community visits. One community member shared ‘When I took my baby, I was nervous. I spoke to the nurses at the health centre… They explained what the vaccine would do. Even though my mind was worried, what they explained helped’ FGD 009. The health information provided in local languages (e.g., Kaqchikel) from multilingual health workers improved trust and understanding in Indigenous communities. One community member said ‘at the health centre, even if you speak Kaqchikel, there are nurses there who share that language… The nurses help you get information’ FGD 001. Another community member said:

> ‘In the villages, they still have a lot of people that don’t understand Spanish…. what they have done in the health centre is they explain it in Spanish first and the translator goes in Kaqchikel… at the health centre… they explain in two languages’ FGD 002.

### Changing vaccine attitudes

Several community members expressed a belief that vaccine confidence was becoming a norm in their communities. Most community members reported vaccinating their own children. Several indicated they had vaccinated or would vaccinate their children with the HPV vaccine. Responses reflected an idea that vaccine attitudes were changing and there was a belief that ‘now viruses are prevented through vaccines’ FGD 007. One community member shared:

> ‘I was never vaccinated… Not like children now… I caught whooping cough… Now, if we didn’t vaccinate our children, the same thing would happen. But thank God, because we have these vaccinations, they don’t get these diseases anymore’ FGD 002.

Another community member shared:

> ‘I have had my daughters vaccinated [against HPV]… at the school they tell them that the vaccine is good. There is no need to be scared because it vaccinates against cancer… it was not for us, we are already the past, but they are the future’ FGD 003.

Community members and health workers indicated a noticeable shift in attitudes towards health, vaccines, society, and communication across different generations. A conversation between three community members illustrates this experience:

> Community member 1: ‘If my two children come up to me and ask, “Mom, how does this work?…” I sit down with them and I say that human papilloma is transmitted through sexual intercourse… I stress the importance of protection during sexual activity and being selective about partners. It’s crucial to understand that the vaccine doesn’t provide a free pass for unrestricted sexual activity. I openly discuss these matters with my daughters… Nobody told me anything. Even for basic things, like my first period, I had to figure it out by observing my sisters’.

> Community member 2: ‘My grandmother used to tell me that if I kissed a boy, I would get pregnant… Now, when I talk to my daughters, I caution them about kissing but not about becoming pregnant from it…

> Community member 3: ‘When I was growing up I didn’t understand everything well… In my house, they didn’t talk like that to the children… My first period… They never talk to me about that… Now it is very different. I try to talk openly with my daughter so that she is well-informed about what’s happening, unlike the past when children were kept in the dark…’ FGD 006.

Similarly, many people expressed that vaccine hesitancy was more common in older generations and less common in younger generations. One community member shared ‘My grandparents were unwilling to vaccinate. They also didn’t understand about vaccines and fevers, so they were afraid of vaccines’ FGD 007. A health worker said ‘[their grandparents] thought that since they grew up without any vaccines, their children would also be fine without vaccines… They didn’t see the benefit of anything’ IDI 008. Another community member said ‘In the past, grandparents viewed vaccines as something bad for children… Now, it’s uncommon for children to solely rely on herbs for their health. In today’s context, all children get vaccinated’ FGD 006.

Another health worker said:

> ‘In the past, grandparents were less likely to get vaccinated. They didn’t go to a health centre, a health post, or a hospital. They relied on midwives, mountains, and natural waters. They don’t believe in chemical interventions’ IDI 004.

They expressed that the difference is partly because, unlike their elders who leaned towards traditional medicine and faced limited access to healthcare, younger people are more open to modern technologies, information, and medical practices, including vaccinations. Two community members discussed this:

> Community member 1: ‘I have seen these things with my children… with the Internet, they have all the information at their fingertips… we may not fully understand how to use the Internet, but our children are more advanced than us.’ Community member 2: ‘She already knows. Maybe you say, “Oh, she’s not old enough to discuss such things, to be a woman.”… they already have knowledge… it’s not like before … Now, with the internet… there are things they understand better than we do’ FGD 006.

Health workers shared this idea, with one stating ‘nowadays, I think the youth is a little bit more informed and more accepting of vaccination’ IDI 015. Another health worker said ‘we already have an open-minded population, mothers are concerned about the welfare of their daughters nowadays because they talk about the consequences later on’ IDI 019. Another health worker confirmed this idea stating:

> The community respects our parents’ beliefs. However, the decision ultimately lies with each individual based on their experiences, knowledge, or studies. Younger people tend to be more easily convinced [of the benefits of vaccination] than older individuals’ IDI 018.

### Social networks and community pressure to vaccinate

Both community members and health workers highlighted the role of family members and community networks in sharing health information and encouraging or discouraging vaccination. One community member said ‘I always share everything. I call my siblings to see if my nephews were vaccinated or not. They tell me. I start by investigating whether they did or not’ FGD 008. A health worker stated ‘well, some who do not have access to health centres rely on those who go to the centres or hospitals the most to bring them information. Let’s say a family member… they’re telling each other’ IDI 012.

Community members noted that mothers faced pressure from family and community to vaccinate or not vaccinate their children. If a child became ill, mothers were often judged. Of note, mothers could be judged if others believed she provided inadequate care for the child after the vaccination. Two community members discussed this stating:

> Community member 1: ‘Mothers feel a lot of responsibility because if something happens to the children, they are the ones to blame… There may be a level of machismo that still exists because I still hear cases where they say… your daughter is sick because you gave her a vaccine’.

> Community member 2: ‘Yes, what the mother says is true… Several mothers did not want to vaccinate their babies because they said that their husbands scolded them. They have forbidden them to vaccinate their children. That’s why they have this fear. That’s why they don’t vaccinate their children’ FGD 001.

When asked about perceived vaccine risks two community members shared:

> Interviewer: ‘So, there are children who have died [from being vaccinated]?’

> Community member 1: ‘Yes, there are children, but it’s more about the mother’s care. Yes, it’s the care’.

> Community member 2: ‘It’s not the vaccine, it’s the care… it depends on the care of a mother’ FGD 003.

Community members and health workers identified local leaders, such as midwives, teachers, religious leaders, and other community figures, as trusted sources of vaccine information. They noted that announcements from government health authorities and health centres, particularly when endorsed by community leaders or delivered through formal channels, were trusted sources of information. One community member shared ‘at the schools, one of the health centres comes to vaccinate our children. There the teachers give us a message a day before so that the children will get vaccinated’ FGD 007. Health workers agreed, sharing ‘the Ministry of Health, and also the community leaders, for example, if the midwives go and say, ah, yes the HPV vaccine is important to prevent womb cancer, then the women are going to trust it’ IDI 007. Another health worker said ‘people say that if the pastor of the church said it, it is because it is true. If the community assistant says it, then if it is true. People are more likely to believe information coming from these trusted community figures’ IDI 018.

## Discussion

Childhood immunisation and HPV vaccines have been on the decline in Guatemala since before the COVID-19 pandemic [1]. The present study sought to examine barriers to vaccination and health information needs in three departments in Guatemala. Indigenous community members expressed fears about vaccines, often due to limited information on their purpose, need, and potential side effects. Significantly limited knowledge about HPV and the HPV vaccine was apparent. These barriers are aligned with previous research in Guatemala [11–15]. Systemic issues including lack of knowledge, skills, and training affected health workers, increasing community distrust, though local health workers remained a key source of information, especially in Indigenous Mayan languages.

In the absence of information, rumours about infertility or paralysis linked to vaccines spread through social networks, causing doubts despite some hesitancy to believe them. These networks also pressured parents, especially mothers, to ensure their children’s health, which could either hinder or support vaccination depending on individual beliefs within the social network.

Other systemic barriers, such as practical access issues, were less frequently mentioned but included temporary vaccine shortages at public centres, forcing families to buy vaccines at high costs, and long lines at health centres and clinics. Other Guatemalan studies have indicated practical access issues as being significant [20]. Practical access may have been less of a concern due to the communities’ connection with Wuqu’ Kawoq, which may provide better access to health services and vaccines. Alternatively, the study’s focus on communication barriers for a vaccine uptake campaign may have led participants to prioritize those over structural issues. More research should be done on these potential barriers to ensure that our findings hold.

Even though community members and health workers identified vaccine barriers they expressed a fervent belief that attitudes toward health and society were changing, as well as new access to health, information, and technology was emerging. Many interviewed were positive that these changes would contribute to improved health and vaccination rates. This suggests that there is an opportunity to further change attitudes and behaviours and leverage new communication technologies.

### Limitations

Given the sample was restricted to eleven communities in the Central Highlands of Guatemala, results cannot necessarily be generalised to the wider population, or even other Indigenous groups. Those who chose to participate in focus group discussions about vaccinations may be systematically different from those who did not agree to participate. Social desirability bias may have also impacted the responses during the interviews and the focus groups.

## Conclusions

This study sheds light on the multifaceted barriers to childhood immunisations and HPV vaccination within rural Indigenous communities in Guatemala. By identifying individual, interpersonal, and systemic barriers it underscores the complexity of vaccination uptake in these settings. The reliance on social networks for health information underscores the importance of targeted communication strategies. The growth of social networks over social media could potentially offer an impactful way of addressing attitudes and behaviours through social norm change and increased information in this setting. While practical access issues emerged as less prominent, they should be considered for further research, especially in the context of designing vaccination campaigns.

Despite challenges to vaccine uptake, community members and health workers expressed optimism regarding potential attitude shifts and enhanced access to health information and technology to improve vaccination rates. However, the study acknowledges its limitations, particularly the restricted sample size and the potential for bias in participant selection and responses. Moving forward, broader research efforts are needed to validate these findings across Guatemala’s Indigenous populations and to develop tailored interventions that address the unique needs of Indigenous communities in Guatemala and beyond.

## Authors’ Contributions

All authors contributed to the design of the study including research questions and instruments. MS, JC, LAM, and EL led the interviews, focus groups, recruitment, and translations. NAS, LAM, EL, and JSJ led the coding and analysis. NAS wrote initial manuscript drafts. JSJ, LAM, NDS, EL, and AKD revised the manuscript. All authors reviewed the final drafts of the manuscript.

## Acknowledgments

We would like to thank the community members who participated in focus groups for their generous contributions to this project by sharing their lived experiences and personal opinions. We would also like to thank the health workers who shared their experiences with vaccine campaigns in Guatemala and contributed ideas on how to improve vaccine communications in their communities. We would like to thank the entire Wuqu’ Kawoq staff, including their participation in interviews and connections to both local health workers and community members. We would also like to extend our gratitude to the entire Institute for Global Health Sciences at the University of California, San Francisco group, especially Esbeydy Pardo, the Stanford Center for Health Education Digital Medic team including Semay Johnston, Katherine Sziraczky, Aarti Porwal, and Charles Prober, and Creative Frontiers.

## Ethics Statement

Verbal informed consent was obtained from the study participants. All participants were read the consent document in private to ensure that regardless of literacy they were fully informed about their right to refuse participation and to ensure they were clear on the purposes of the research project. This study has been approved by the Institutional Review Boards at Stanford University (Protocol # 63193), the University of California, San Francisco (Study #23-39043), and a private IRB through Wuqu’ Kawoq (Protocol # WK 2024 003). No members of the research team have financial interests in the development of vaccines or vaccine promotion.

## Data Availability Statement

No data are available.

